# A Fusion-Based Multiomics Classification Approach for Enhanced Gene Discovery in Non-Small Cell Lung Cancer

**DOI:** 10.1101/2025.05.02.25326847

**Authors:** Kountay Dwivedi, Amirreza Mahbod, Rupert C. Ecker, Klara Janjić

## Abstract

This study introduces a fusion-based multiomics approach to identifying non-small cell lung cancer (NSCLC)-relevant genes. We evaluated the NSCLC-subtype classification performance of various state-of-the-art machine learning models using single omics and fused multiomics approaches. The models were trained separately on individual omics data sets. Subsequently, a weighted-average-based decision-level fusion mechanism was employed to integrate the individual predictions of the trained models. Finally, the prediction performance across all the approaches was compared. The decision-level fusion-based approach yielded a superior classification performance as compared to the performance achieved by models trained on individual omics data sets. Finally, a set of 47 NSCLC-relevant genes were identified. For the first time, *ABCF3*, *ACAP2*, *LSG1*, *TBCCD1*, *UCN2*, *WDR53*, *ZNF639* and *FYTTD1* appeared in the context of NSCLC. In conclusion, the integration of multiple omics types showed potential to deliver a more concise selection of NSCLC-relevant genes that could be clinically targeted in future.

**Graphical abstract:** 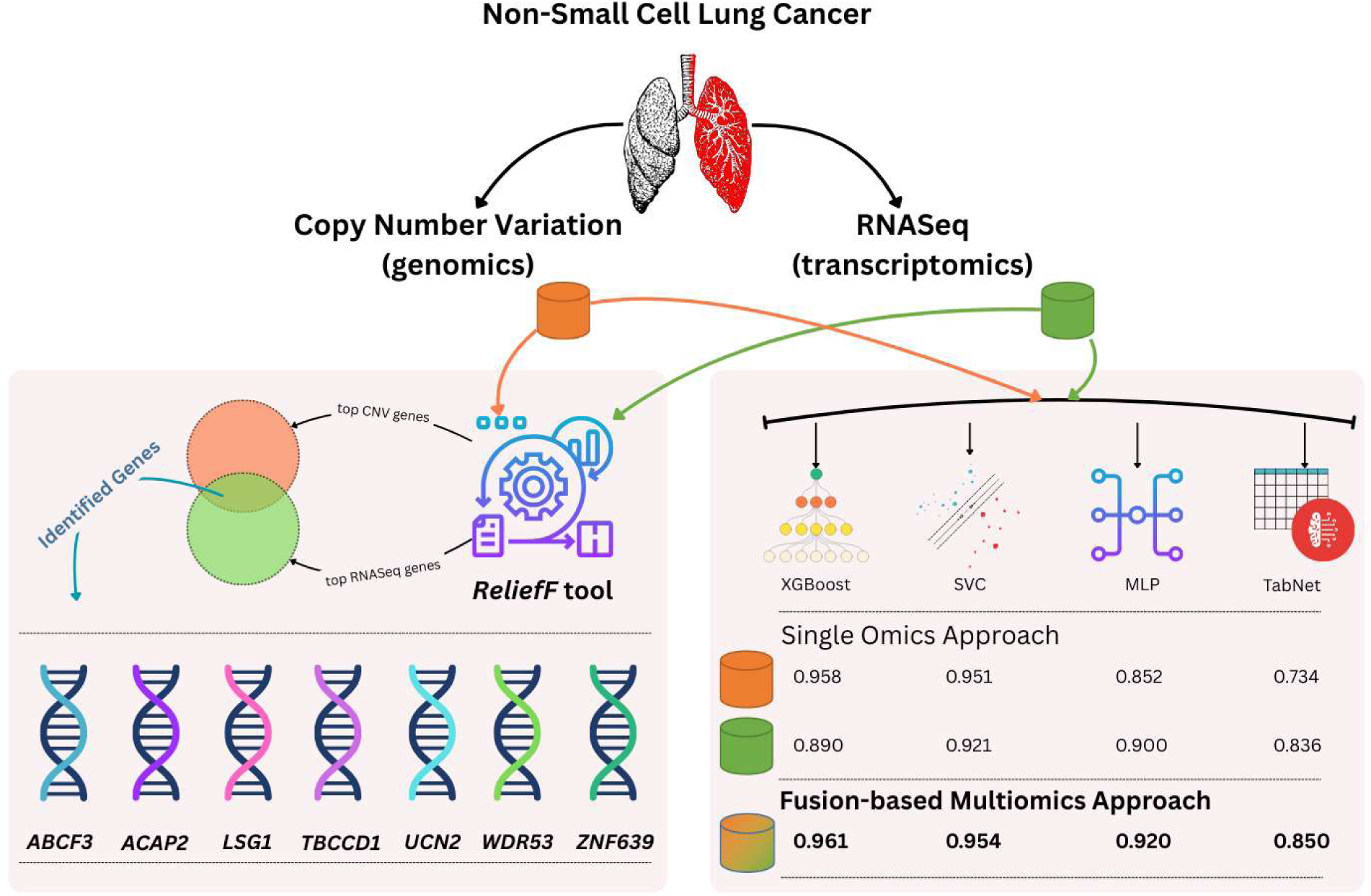

## 1 Introduction

Over the years, lung cancer has emerged as the most prevalent cancer worldwide, showing incremental growth in incidents and mortality, while depicting a 5-year survival rate of approximately 17% (Siegel *et al*., 2022; Dwivedi *et al*., 2023). Non-small cell lung cancer (NSCLC), being the most prominent lung cancer type, represents around 85% of the cases (Dwivedi *et al*., 2023). The advancements in molecular pathology further aided in the pathological-level subtype classification of NSCLC as adenocarcinoma (ADC) and squamous cell carcinoma (SCC) (Travis *et al*., 2015). This sub-categorization is based on the distinct and heterogeneous pathogenic characteristics portrayed by a characteristic gene expression profile of the tumor cells (Guinney *et al*., 2015; Travis *et al*., 2015; Chen *et al*., 2022; Lipkova *et al*., 2022; Yang *et al*., 2022). It is, thus, imperative to identify the key genes accountable for the alterations leading to tumor development, subsequently deriving potential therapeutic targets (Travis *et al*., 2015; Girard *et al*., 2016; Chen and Dhahbi, 2021; Dwivedi *et al*., 2023).

Throughout literature, several studies have contributed to shaping the landscape of NSCLC genes identification and subtype prediction by investigating omics-level data, such as genomics, transcriptomics and epigenomics (Li *et al*., 2014; Cai *et al*., 2015; Girard *et al*., 2016; Qiu *et al*., 2017; Tian, 2017; Chen and Dhahbi, 2021; Dwivedi *et al*., 2023, 2024a, 2024b). A notable work employing transcriptomics data was published by Girard *et al*. (Girard *et al*., 2016) who employed RNASeq and microarray-based gene expression data by utilizing volcano plots and Pearson’s correlation to identify 42 genes with an accuracy of 0.950. Chen and Dhahbi (Chen and Dhahbi, 2021) applied multiple feature selection methods to identify 17 relevant transcriptomics genes. By employing a random forest model (Breiman, 2001), they achieved an accuracy of 0.929. Tian (Tian, 2017) developed a pipeline integrating GeneRanks (Morrison *et al*., 2005) and RadViz (Hoffman *et al*., 1997) projection methods to rank the transcriptomics genes. From this ranking, the author identified 8 genes that aided in achieving a classification accuracy of 0.924 via a support vector classifier model (Cortes and Vapnik, 1995). Recently, Dwivedi *et al*. (Dwivedi *et al*., 2023) leveraged deep learning and SHapley Additive exPlanations (SHAP)-based interpretability technique (Shapley and others, 1953; Lundberg and Lee, 2017) to identify a set of 52 genes with a prediction accuracy of 0.957.

Another essential omics type, the DNA copy number variation (CNV) acts as a key attribute in human genome development (Pinkel and Albertson, 2005; Pfarr *et al*., 2016). The CNV information from the cancer genome may assist in identifying recurrent chromosomal alterations that, in turn, could aid in targeting cancer-related genes (Kim *et al*., 2013). Qiu *et al*. (Qiu *et al*., 2017) developed a custom classifier to identify a panel of 33 NSCLC-relevant CNV genes for classification. Using the identified genes as a feature set, the authors achieved an accuracy of 0.840. Li *et al*. (Li *et al*., 2014) leveraged minimal redundancy maximum relevance (Peng, Long and Ding, 2005) algorithm to rank the CNV probes, thereafter utilized k-nearest neighbor (Fix and Hodges, 1989) and incremental feature selection (Liu and Setiono, 1998) algorithms to select a set of 266 CNV probes that yielded the best predictive score of 0.860 when validated using leave-one-out cross-validation method.

Based on the aforementioned studies, it can be implied that individual omics data can sufficiently identify a set of NSCLC-relevant genes. Nonetheless, an intricate disease such as NSCLC exhibits various underlying interdependent biological characteristics that can be better interpreted by following a multiomics approach (Yang *et al*., 2022; Khadirnaikar, Shukla and Prasanna, 2023; Steyaert *et al*., 2023). By fairly integrating a diverse set of omics data, a more sensitive and complementary set of genes can be identified that can be used subsequently to improve targeted therapy (Carrillo-Perez *et al*., 2022; Yang *et al*., 2022). This task can be accomplished by utilizing a decision-level fusion method such as hard/soft -voting scheme and weighted-average mechanism (Rastghalam and Pourghassem, 2016; Qi *et al*., 2021; Carrillo-Perez *et al*., 2022). In these approaches, the outputs of isolated omics data are integrated or “fused” to attain a more stable and robust result. Following the decision-level concept, Carrillo Perez *et al*. (Carrillo-Perez *et al*., 2022) utilized a weight-sum optimization method to fuse prediction probabilities obtained by analyzing transcriptomics, genomics, epigenomics as well as histological data of lung cancer. With the proposed method, the authors achieved a prediction accuracy of 0.955.

Here, we assert that, while an omics data used in isolation can identify relevant genes, its efficacy is limited by the inability to adequately capture the inter/intra -biological dependencies. In light of this, the present work proposes the identification of key genes for NSCLC by integrating data from diverse omics types, particularly transcriptomics (RNASeq gene expression) and genomics (CNV). To this end, we present a comprehensive benchmark study evaluating the classification performance of diverse, cutting-edge machine learning (ML) models trained on the aforementioned types of omics data sets, both individually and in a fused manner. We argue that the samples belonging to either of the NSCLC subtypes would be classified more accurately when the predictions derived from the two omics data sets are fused at the decision-level rather than used independently. The fusion approach enhances the model’s ability to capture complex interconnected dependencies among features. Consequently, the most influential features contributing towards the classification could, therefore, be discovered and further examined for their clinical relevance in the diagnosis and prognosis of NSCLC.

The aim of this study is to discover a concise set of NSCLC-relevant genes using a multiomics approach. For this purpose, we evaluated the classification performance of diverse ML models, including extreme gradient boosting (XGB) (Chen and Guestrin, 2016), support vector classifier (SVC) (Cortes and Vapnik, 1995), multilayer perceptron (MLP) (Haykin, 1994) and a transformer-based deep learning model TabNet (Arik and Pfister, 2021) on publicly available ADC and SCC cohorts generated by The Cancer Genome Atlas (TCGA) program, comparing the performance based on individual as well as fused omics data sets. Finally, the fused omics data set is leveraged to identify a small set of NSCLC-relevant genes found most crucial in lung cancer classification.

## 2 Methods

### 2.1 Data Set and Preprocessing

The experimentation was performed on publicly available data sets generated by the TCGA program. The RNASeq and the CNV data sets were downloaded from the UCSC Xena (Goldman *et al*., 2020a) repository on December 27, 2024. The number of samples per omics type for ADC and SCC is mentioned in **Table 1**, while the demographic details of the cohorts are summarized in **Figure 1**.

**Figure 1.**
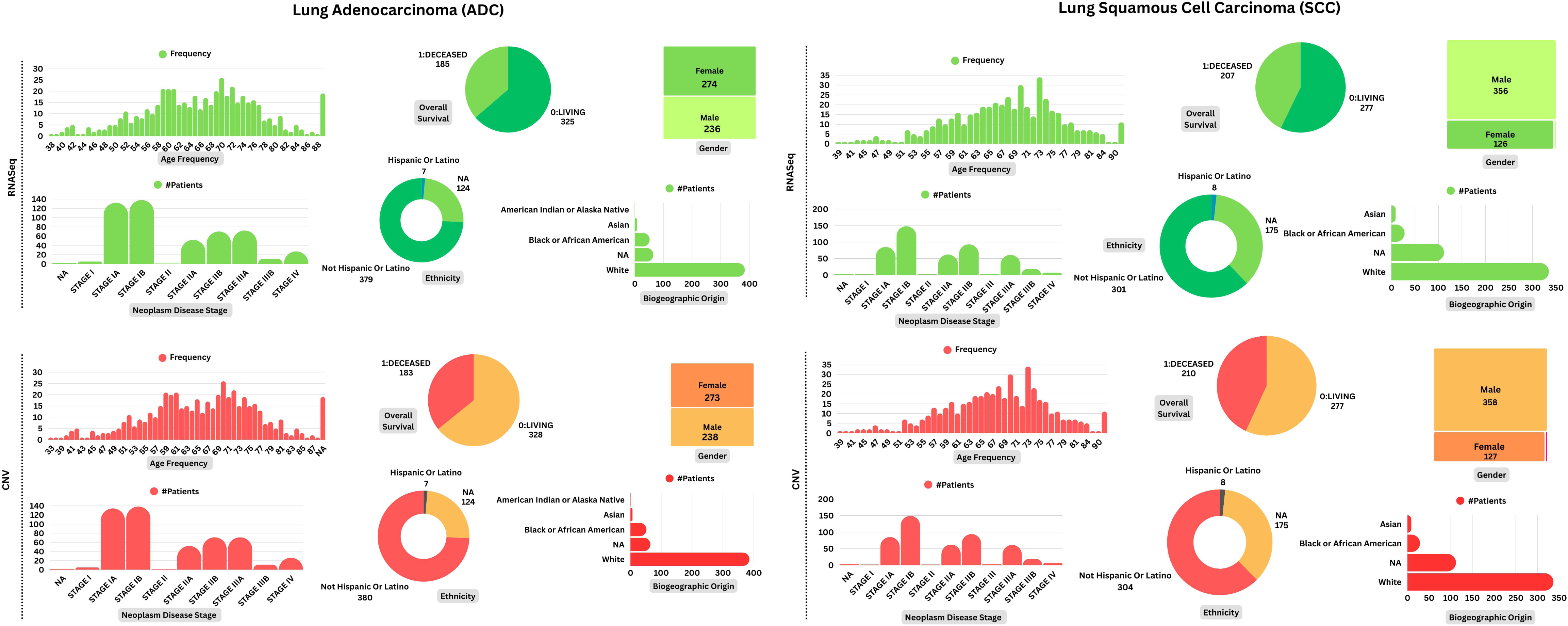
The demographic details of adenocarcinoma and squamous cell carcinoma patients falling under individual omics type. In the transcriptomics data set, the demographic data was available for 510 out of 576 adenocarcinoma (ADC) patients and 484 out of 553 squamous cell carcinoma (SCC) patients. Similarly, in the genomics data set, the demographic information was present for 511 out of 516 ADC patients and 487 out of 501 SCC patients.

**Table 1.** The number of samples and genes per omics type. An effective execution of the decision-level fusion mechanism necessitates the selection of samples with both types of omics data in the non-small cell lung cancer subtypes lung adenocarcinoma (ADC) and squamous cell carcinoma (SCC).

|  | ADC | SCC | Genes |
| --- | --- | --- | --- |
| RNASeq Gene Expression | 576 | 553 | 20,530 |
| Copy Number Variation | 516 | 501 | 24,776 |
| **Common samples** | **505** | **483** |  |

#### 2.1.1 Omics Data Generation Process

The UCSC Xena repository provided the process of generating various omics-level data (Goldman *et al*., 2020b). The RNASeq-based mRNA analysis performed to compute the expression level of individual genes in each sample under study was initiated by obtaining raw read counts aligned against the GRCh38 reference genome using the STAR software (Dobin *et al*., 2013) via HiSeq Illumina sequencers. Subsequently, the augmentation of read counts was performed using various transformation methods such as transcripts per million (TPM), fragments per kilobase of transcript per million mapped reads (FPKM) and upper quartile normalized FPKM (FPKM-UQ). The process was concluded by annotating the augmented read values with the gene symbols and gene bio-type (Dobin *et al*., 2013), thereafter, computing the log_2_(*count* + 1) normalization of the read counts.

The CNV was estimated at the gene level by employing the GISTIC2 methodology (Mermel *et al*., 2011). Initially, the raw copy number values were generated using the Affymetrix Single Nucleotide Polymorphism (SNP) 6.0 platform at the Broad Institute as part of the TCGA project. Subsequently, the obtained values were log_2_ *x*-normalized and segmented using the circular binary segmentation (CBS) (Olshen *et al*., 2004) algorithm. These segmented copy number values were conclusively mapped to the associated genes, resulting in gene-level copy number estimates. The segment-to-gene mapping was performed using the UCSC Xena HUGO probeMap (Mermel *et al*., 2011).

#### 2.1.2 Data Set Preprocessing

To implement an effective decision-level fusion mechanism, it was necessary to use the samples that contain both omics data. Accordingly, each omics data set was initially examined to identify common samples across modalities, yielding a set of 505 and 483 samples in ADC and SCC data sets, respectively as shown in **Table 1**. Subsequently, when the data sets were investigated for missing values, no missing data were found. Finally, each data set was z-score normalized to scale its mean and standard deviation to zero and one, respectively.

### 2.2 Model Training, Validation and Decision-level Fusion

For the purpose of training and validation, the state-of-the-art ML models were employed, including XGB, SVC, MLP and the transformer-based TabNet model. All the models were utilized with their default hyperparameter settings. Initially, each model was trained independently on individual omics data sets. Subsequently, the resulting predictive probabilities obtained from each omics data set were integrated using a *weighted-averaging* technique to compute the integrated or “*fused*” predictive probability.

To cater the inherent stochastic nature of the computing system, the aforementioned process was iterated 10 times – each time using a different random seed. For each iteration, the models were trained and validated using a 5-fold stratified cross-validation approach to ensure a robust model selection.

#### 2.2.1 Weighted-Average-based Decision-Level Fusion Mechanism

To ensure the accurate integration of the predictive probabilities computed from individual models, the same samples were maintained in the train and validation splits across all the omics data sets. Formally, for NSCLC subtype prediction – a binary classification problem – consider a set of validation samples *m*, each with an associated class label *y* ∈ {0, 1} ∀*m*. Then, for each omics-specific model *F_i_*: *x* → *y* where *i* ∈ {RNASeq, CNV} and *x* ∈ *m*, the output is a pair of prediction probabilities {*P_F_i__* (*y* = 0), *P_F_i__* (*y* = 1)} that sum up to 1 (Karlos, Kostopoulos and Kotsiantis, 2020). Let *acc* be the accuracy computed by *F_i_*, then:

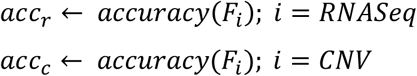

The weighted-average-based decision-level fusion mechanism integrates or *“fuses”* the predictive probability for each *y* and finally outputs the decision-level fusion prediction *ỹ* as:

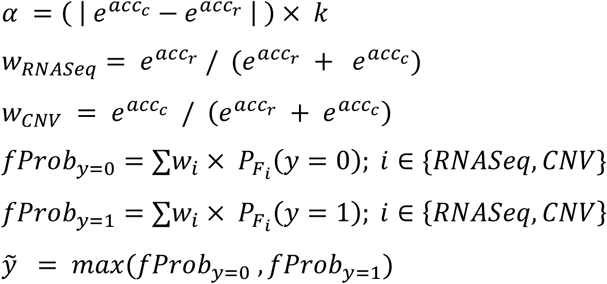

where *k* is the scaling factor (determined experimentally).

### 2.3 ReliefF-based Predictive Genes Identification

In order to identify a concise set of genes most crucial in NSCLC subtype prediction, we used the *ReliefF* (Kononenko, Šimec and Robnik-Šikonja, 1997) method, implemented in the *scikit-rebate* (Urbanowicz *et al*., 2018) library. As highlighted by Bolon-Canedo *et al*. (Bolón-Canedo, Sánchez-Maroño and Alonso-Betanzos, 2013) and Urbanowicz *et al*. (Urbanowicz *et al*., 2018), *Relief*-based algorithms are robust feature selection algorithms highly capable of learning complex feature dependencies, such as gene-gene interactions, in predictive modeling tasks. The original Relief method estimates the importance of an attribute based on how distinct its value is among the samples near to each other (Kononenko, 1994). Assume there is a sample *X* belonging to a class *Y* ∈ {0, 1} with a set of attributes with cardinality *M*. Relief initially searches two nearest neighbors of *X* −− *Hit* belonging to the same class as *X* (called *nearest hit*) and *Miss* belonging to the other remaining class (called *nearest miss*).

Assume *n* to be a subset of randomly selected samples, the weights initialization takes places as presented in Algorithm 1:

#### Algorithm 1

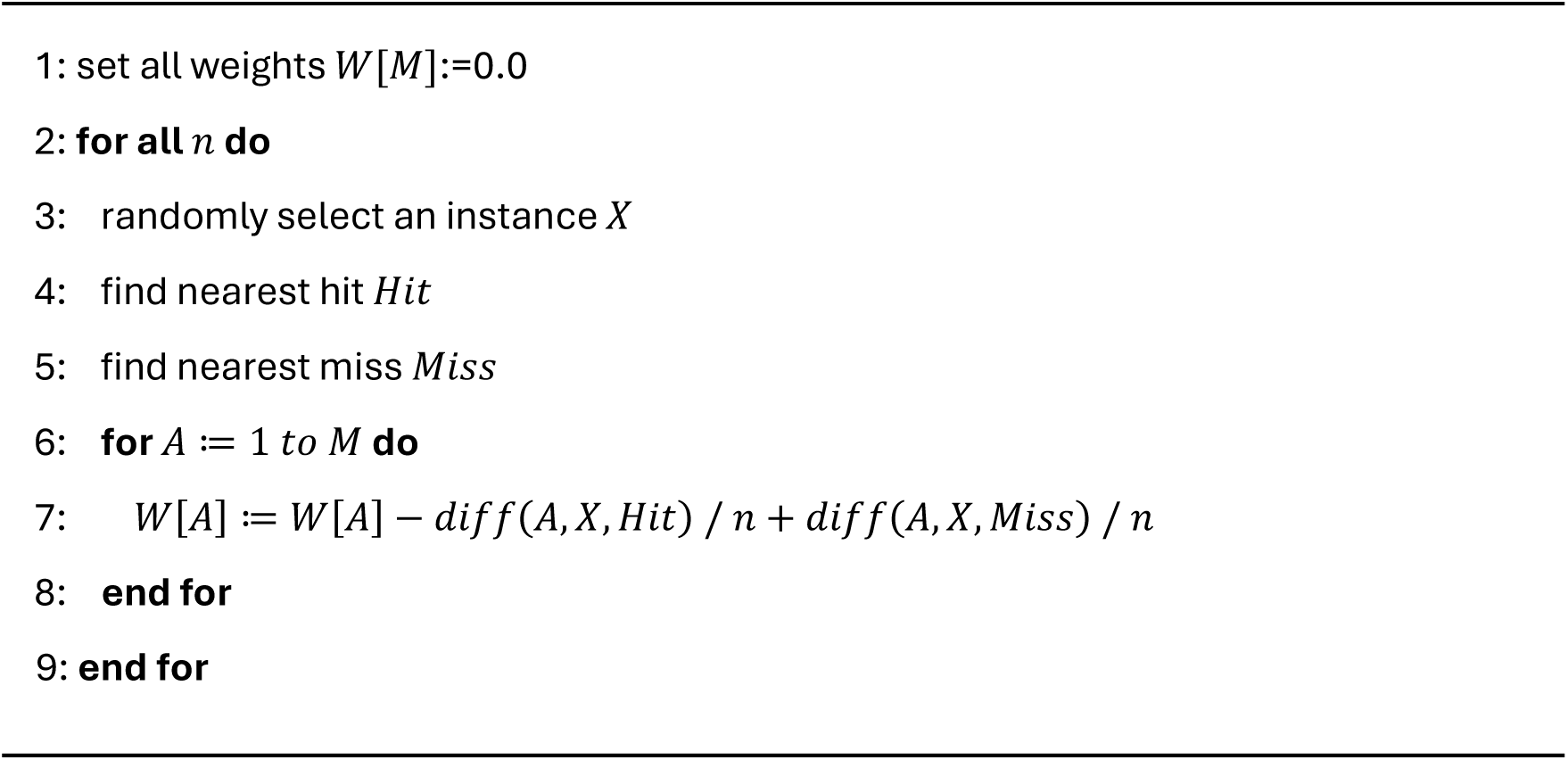

Unlike the original *Relief* method, the *ReliefF* searches for *k nearest hits/misses* instead of just one, thereby increasing the reliability of the resultant weights-approximation (Kononenko, Šimec and Robnik-Šikonja, 1997). To identify a small set of key genes for accurate NSCLC subtype prediction, the *ReliefF* method was initially utilized to compute the importance weights of the genes (attributes) within each individual omics data set – transcriptomics and genomics. Following the computation of the weights, the genes in each omics data set were ranked based on their respective importance scores. Subsequently, the ranked gene lists from both types of omics data sets were intersected to extract a small subset of most influential genes commonly identified as important across both data sets. This intersection represented a set of the most significant predictive genes for NSCLC.

### 2.4 Functional Enrichment Analysis of the Identified Genes

Having discovered the crucial subset of NSCLC-relevant genes, we proceeded to investigate their biological relevance. Initially, we explored the functional enrichment of the discovered set of genes. This analysis was performed in terms of gene ontology (GO) and biological pathways by utilizing the online available WEB-based GEne SeT AnaLysis Toolkit (WebGestalt) (Zhang, Kirov and Snoddy, 2005). The databases utilized to analyze the functional pathways were Protein ANalysis THrough Evolutionary Relationships (PANTHER) (Mi and Thomas, 2009; Thomas *et al*., 2022), Kyoto Encyclopedia of Genes and Genomes (KEGG) (Kanehisa and Goto, 2000) and Reactome (Fabregat *et al*., 2017).

### 2.5 Overall Survival-based Prognostic Efficacy of the Genes

Post-investigating the functional and biological enrichment of the discovered genes, we investigated their survival prediction performance. The evaluation of the prognostic efficacy of the discovered set of genes was carried out by utilizing the online Kaplan-Meier (KM) Plotter (Györffy *et al*., 2010) tool, which provides transcriptome profiles of large sample sets from multiple cancer cohorts and performs real-time survival analysis by employing Cox regression and KM plots (Lánczky and Györffy, 2021). For this analysis, the lung cancer cohort from the KM Plotter database, comprising 1411 patients, was utilized. The discovered set of 47 genes was provided as an input and the overall survival was computed using the default parameters set by the platform. The survival time was recorded in months.

## 3 Results

### 3.1 Prediction Performance Comparison of Individual Models with Decision-level Fusion Approach

To accomplish an accurate decision-level fusion mechanism, the ML models were first trained on individual omics data sets. This approach facilitated a comprehensive comparison of the predictive performance of individual omics data with the fusion-based approach. The performance was evaluated using a confusion matrix and the AUROC score. The confusion matrix provided a detailed segregation of the predicted labels compared to the true labels. The matrix comprised four cells:

- **True Positives (TP):** The correctly predicted positive samples.
- **False Positives (FP):** The incorrectly predicted positive samples; also known as *Type-I error*.
- **True Negatives (TN):** The correctly predicted negative samples.
- **False Negatives (FN):** The incorrectly predicted negative samples; also known as *Type-II error*.

With the help of the confusion matrix, various classification metrics could be derived, such as accuracy, precision, recall and F1-score, where:

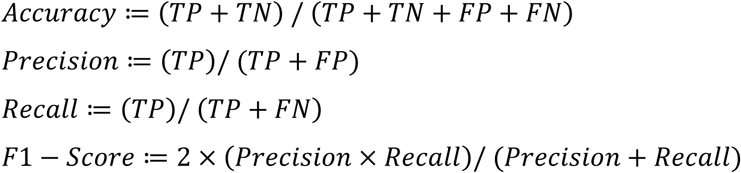

The receiver operator characteristic (ROC) curve in the AUROC plots the True Positive Rate (TPR) against the False Positive Rate (FPR) at various classification thresholds, essentially segregating the “signal” from the “noise”. The area under the ROC curve (AUROC) summarizes the ROC curve by providing the measure of the ability of the model to distinguish between the classes (McClish, 1989). Recall that the models were trained across multiple random seeds to capture the inherent randomness of the system. Therefore, the final confusion matrix and the AUROC curve scores were computed as the average of all the confusion matrices and AUROC curve scores obtained across different random seed iterations.

The comparative analysis of the accuracy, F1-scores and the AUROC scores of different models employed on individual data sets and that of the fusion-based approach revealed that the fusion-based approach outperformed the models trained on the individual data sets (**Table 2**).

**Table 2.** Prediction performance of each individual omics data set alongside fusion-based approach. Both approaches, based on individual omics data as well as the fusion/based approach were performed on four different models, including Extreme Gradient Boosting (XGB), Support Vector Classifier (SVC), Multilayer Perceptron (MLP) and TabNet. The fusion-based approach outperformed the models trained on individual omics data sets, RNASeq (RNA) and copy number variation (CNV) in terms of accuracy, F1-score and the area under the receiver operator characteristic (AUROC) score.

| **Model/Approach** | **Accuracy** | **F1-Score** | **AUROC** |
| --- | --- | --- | --- |
| XGB-RNA | 0.958 | 0.959 | 0.991 |
| XGB-CNV | 0.890 | 0.890 | 0.953 |
| **XGB-Fusion** | **0.961** | **0.961** | **0.982** |
| SVC-RNA | 0.951 | 0.951 | 0.987 |
| SVC-CNV | 0.921 | 0.920 | 0.960 |
| **SVC-Fusion** | **0.954** | **0.954** | **0.980** |
| MLP-RNA | 0.852 | 0.852 | 0.935 |
| MLP-CNV | 0.900 | 0.900 | 0.956 |
| **MLP-Fusion** | **0.920** | **0.920** | **0.971** |
| TabNet-RNA | 0.734 | 0.733 | 0.834 |
| TabNet-CNV | 0.836 | 0.836 | 0.930 |
| **TabNet-Fusion** | **0.850** | **0.848** | **0.931** |

Additionally, we found that the predictive performance across all models was improved in the fusion-based approach compared to the performance based on omics-specific models (**Figure 2**). Further, **Figure 3** includes a line graph illustrating the accuracy and AUROC scores, alongside the corresponding confusion matrices, thus, presenting a comprehensive evaluation of model performance.

**Figure 2.**
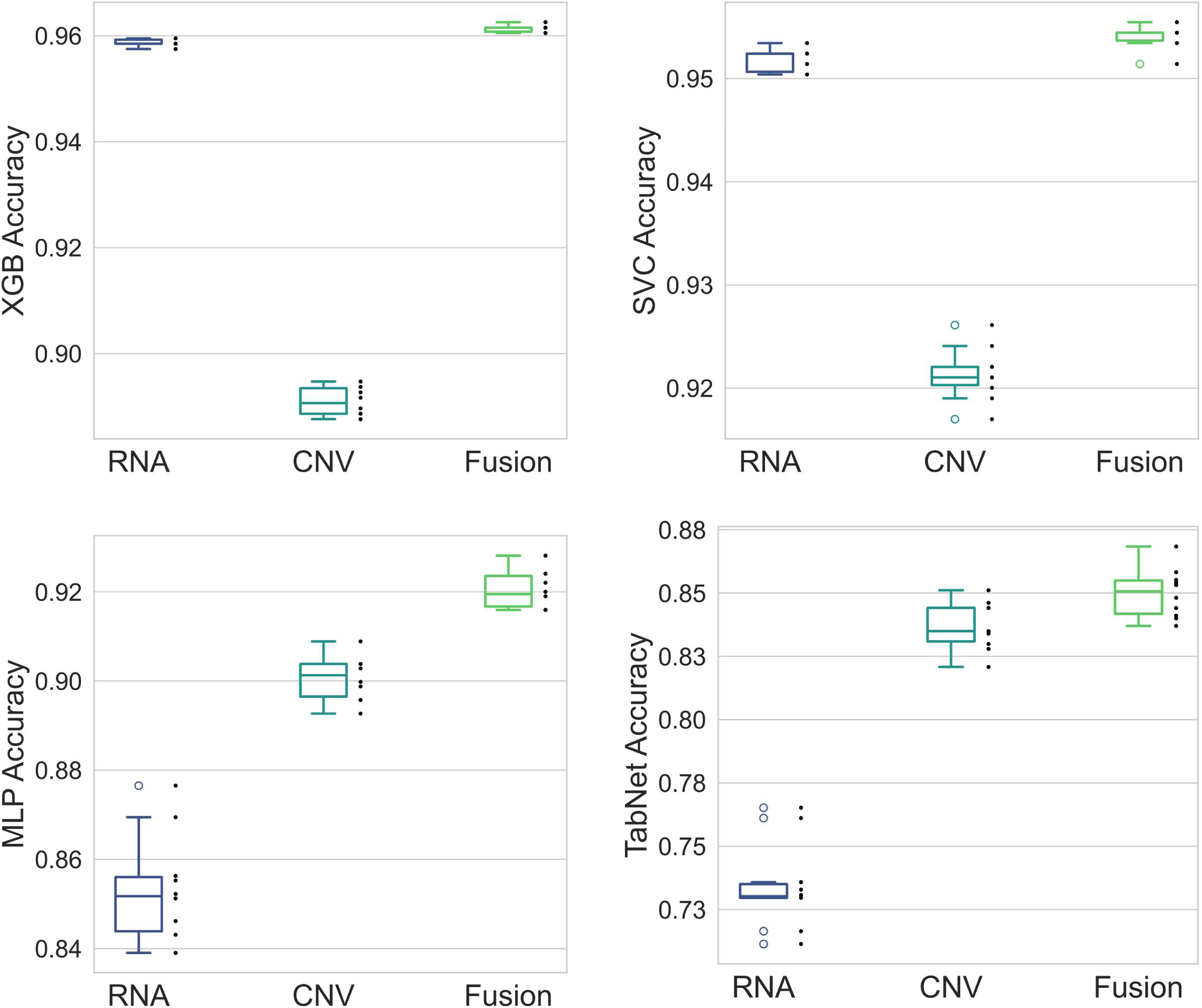
Accuracy achieved by each model when trained on individual omics data set compared to the fusion-based approach. The proposed decision-level fusion mechanism surpasses all the models trained on individual omics data sets, including RNASeq (RNA) and copy number variation (CNV) data. Both approaches were trained on four different models: Extreme Gradient Boosting (XGB), Support Vector Classifier (SVC), Multilayer Perceptron (MLP) and TabNet.

**Figure 3.**
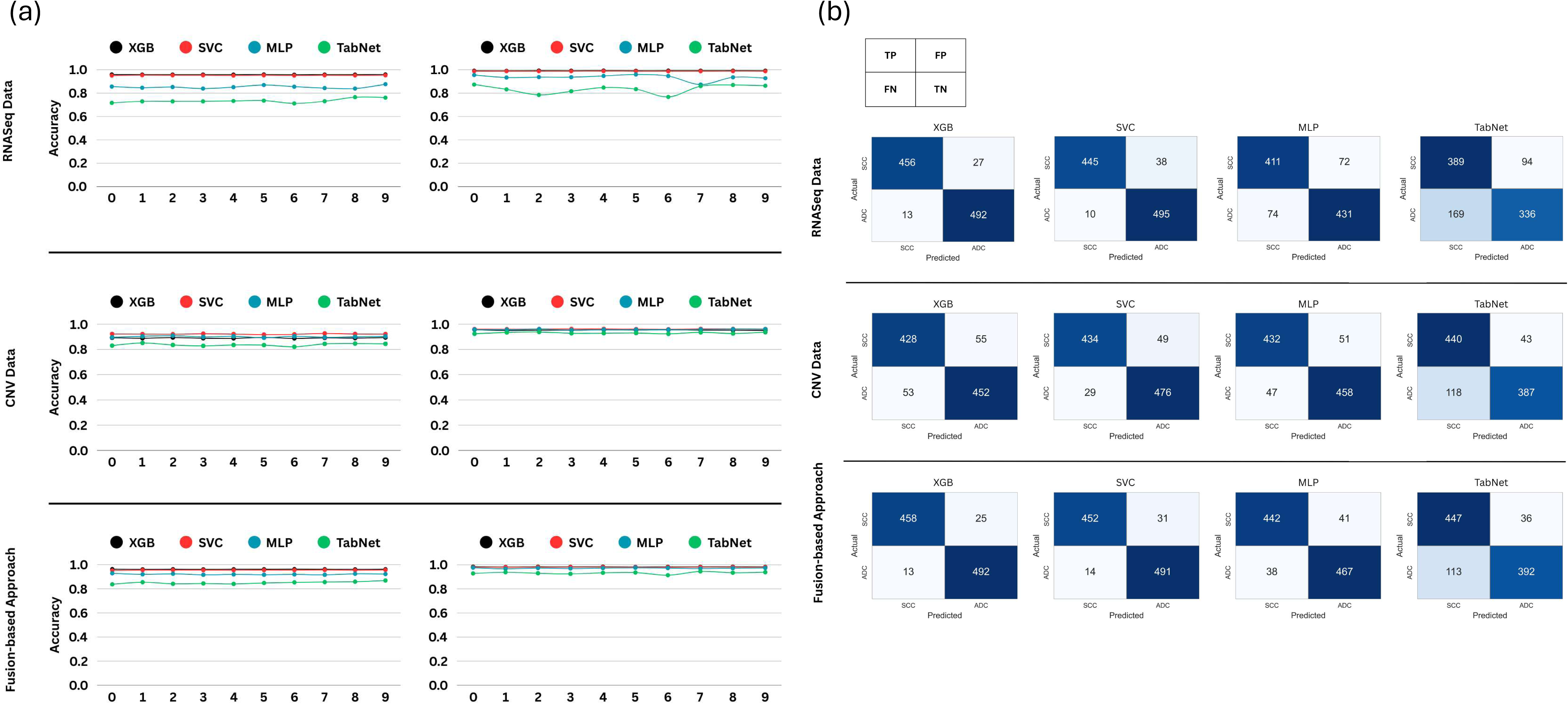
Line graphs and confusion matrices depicting the comparative prediction performance of models when trained on RNASeq and copy number variation data against the performance achieved when the fusion-based approach is employed. In the line graphs (a), the x-axis represents the ten iterations conducted with random seed values, while the y-axis indicates the corresponding performance metrics. The confusion matrices (b) portray the absolute counts and percentages of true positive (TP), false positive (FP), false negative (FN) and true negative (TN) predictions made by each model, respectively, thus providing a comprehensive overview of their classification performance. Notably, the fusion-based approach exhibited superior performance compared to models trained on individual omics datasets, RNASeq (RNA) and copy number variation (CNV) highlighting the advantage of integrating multiomics datasets. This finding applies to all the four utilized models: Extreme Gradient Boosting (XGB), Support Vector Classifier (SVC), Multilayer Perceptron (MLP) and TabNet.

As observed, models trained on the individual omics data sets exhibited substantial variation in their predictive performance (**Table 2** and **Figure 2**). While XGB demonstrated high accuracy for RNASeq data (0.958), it performed worse on CNV data (0.890). A similar result was observed with the SVC model, which showed high accuracy on RNASeq data (0.951) but lower performance on CNV data (0.921). Interestingly, the MLP and TabNet models displayed an opposite outcome. The MLP model achieved an accuracy of 0.852 on RNASeq and performed better on CNV data (0.900). Similarly, the TabNet model achieved an accuracy of 0.734 on RNASeq and better accuracy (0.836) on CNV data.

However, the decision-level fusion strategy demonstrated a consistent superior performance (0.961 using XGB, 0.954 using SVC, 0.920 using MLP and 0.850 using TabNet) compared to all the models trained on individual omics data sets, thus, relatively mitigating the high variation in the models. As an ablation study, we compared our proposed weighted-average-based decision-level fusion mechanism with a standard late fusion employing soft voting mechanism. For this purpose, we utilized the XGBoost model, which demonstrated a superior performance relative to the other employed models. The classification performance was assessed on the basis of accuracy, computed using the class probabilities predicted by XGBoost. The soft voting mechanism yielded an accuracy of 0.958, which was found to be inferior to the performance of our proposed fusion mechanism. Based on these observations, it could be inferred that relying on single omics data may lead to varying predictions that do not identify a concise selection of pathologically relevant key genes.

### 3.2 ReliefF-derived Key NSCLC-relevant Genes

The utility of ReliefF algorithm for gene discovery resulted in an unveiling of a concise set of 47 most relevant NSCLC genes that are consistent across both the omics data sets – transcriptomics and genomics. The complete list of genes is presented in **Figure 4**. It could be confirmed via a literature search that 39 of the identified genes were already associated with lung cancer by either a biological or a computational analysis, which supports the reliability of our fusion-based approach to identify a focused set of key genes. Among the 47 found genes, we discovered a total of 7 genes, namely *ABCF3* (Fan *et al*., 2023), *ACAP2* (Liu *et al*., 2022), *LSG1* (Shen *et al*., 2022), *TBCCD1* (Denu and Burkard, 2020), *UCN2* (Hao *et al*., 2008), *WDR53* (Weissmiller, Fesik and Tansey, 2024) and *ZNF639* (Imoto *et al*., 2003), that could play a role in NSCLC but have only been investigated in the context of other cancer types until now. Another gene, *FYTTD1*, was found as a gene in our analysis which is currently only vaguely studied in pathologies without any indication for its role in any type of cancer yet.

**Figure 4.**
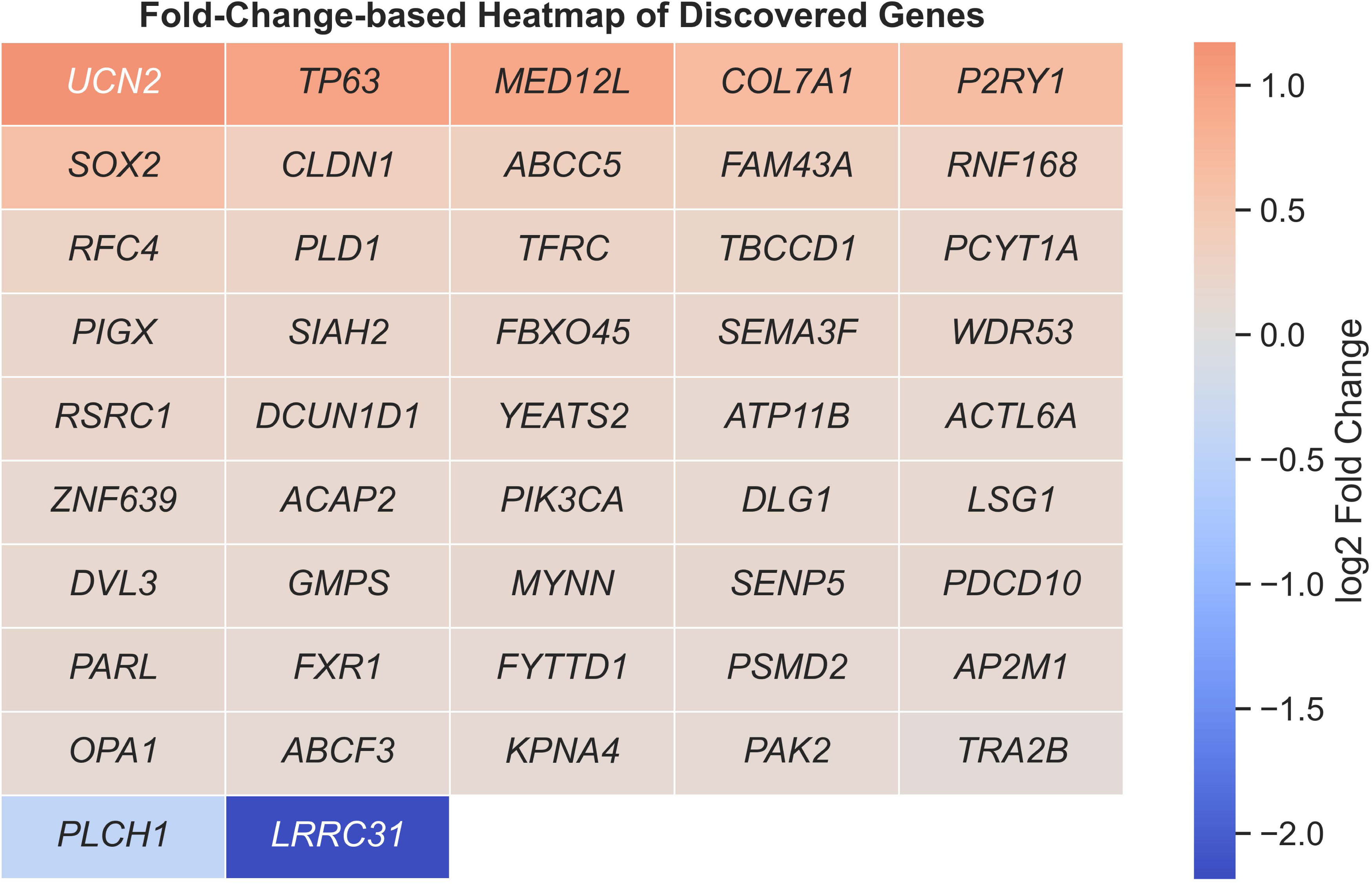
The heatmap corresponds to the log2(*x*) − *based* gene expression fold change of the 47 discovered NSCLC-relevant genes. The two downregulated genes (blue) were *PLCH1* and *LRRC31*, while the highly upregulated genes (red) were *UCN2*, *TP63* and *MED12L*.

### 3.3 Enrichment Analysis of Discovered Genes

In terms of gene ontology (GO) and biological pathways. The discovered genes were found significantly associated with numerous biological processes (**Figure 5**), including *metabolic process, biological regulation, response to stimulus, cellular component organization, localization, multicellular organismal process, cell communication* and *developmental process*. The cellular components found enriched were *membrane, nucleus, protein-containing complex, membrane-enclosed lumen, endomembrane system, cytosol* and *vesicle*. Among the molecular functions, the *protein binding, ion binding, hydrolase activity, nucleotide binding* and *nucleic acid binding* were found enriched.

**Figure 5.**
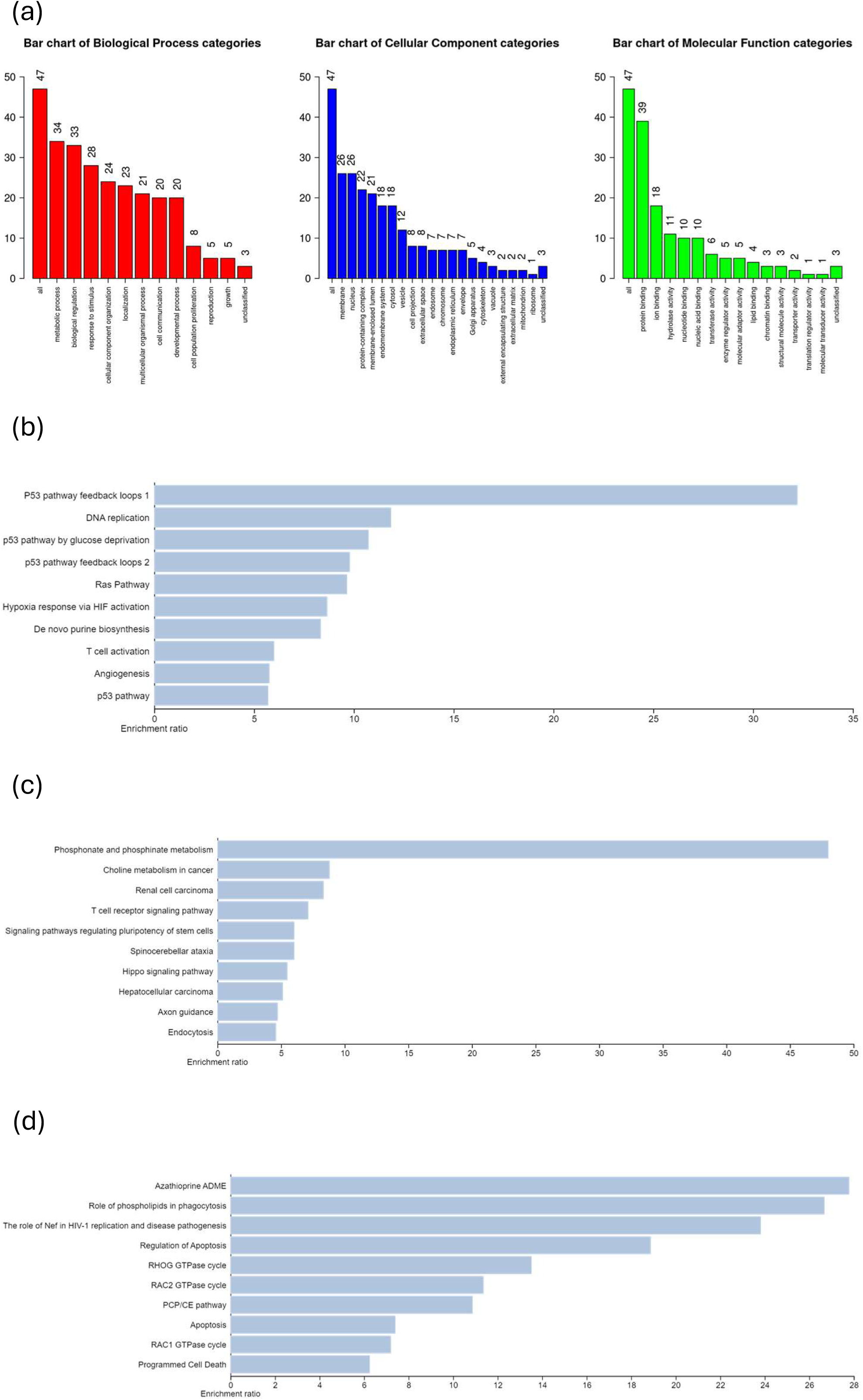
Enrichment analysis of the 47 discovered genes. The discovered genes are associated with a variety of biological processes, cellular components and molecular functions (a). The bar plots illustrate the functional pathways enriched in the 47 discovered genes found in the PANTHER (b), KEGG (c) and Reactome data bases (d), respectively. The x-axis represents the enrichment ratio and the y-axis lists the distinct biological pathways. The enrichment ratio indicates the degree of association of the genes with the respective pathway.

Among the functional pathways, the *Ras pathway* (p=0.002)*, angiogenesis* (p=0.003)*, p53 pathway feedback loops 2* (p=0.016)*, p53 pathway feedback loops 1* (p=0.030)*, T cell activation* (p=0.041) and *p53 pathway* (p=0.045) were found to be significantly enriched in the PANTHER database. The KEGG database showed the *choline metabolism in cancer* (p=0.004)*, T cell receptor signaling pathway* (p=0.008)*, endocytosis* (p=0.0010)*, signaling pathways regulating pluripotency of stem cells* (p=0.012)*, spinocerebellar ataxia* (p=0.012)*, hippo signaling pathway* (p=0.016)*, hepatocellular carcinoma* (p=0.019)*, phosphonate and phosphinate metabolism* (p=0.020)*, renal cell carcinoma* (p=0.023) and *axon guidance* (p=0.024) pathways to be significantly enriched. Finally, the *regulation of apoptosis* (p=0.005)*, RHOG GTPase cycle* (p=0.001)*, apoptosis* (p=0.001)*, RAC1 GTPase cycle* (p=0.002)*, RAC2 GTPase cycle* (p=0.002)*, azathioprine ADME* (p=0.002)*, role of phospholipids in phagocytosis* (p=0.002)*, PCP/CE pathway* (p=0.002)*, the role of Nef in HIV-1 replication and disease pathogenesis* (p=0.003) and *programmed cell death* (p=0.003) were the pathways found enriched in the Reactome database.

### 3.4 Overall Survival Analysis of the Discovered Genes

The evaluation of the prognostic efficacy of the discovered set of 47 genes unveiled a subset of 28 genes that demonstrated significant prognostic potential with p≤0.05. **Figure 6** presents the KM curves of these 28 genes. It is observed that except for *PLCH1 (KIAA1069), LRRC31, RNF168* and *TRA2B*, the lower expression values of the remaining 24 genes were associated with poor prognosis. To further validate this observation, we investigated the combined expression levels of *PLCH1 (KIAA1069), LRRC31, RNF168* and *TRA2B* in healthy and tumor samples using data from TCGA and Genotype-Tissue Expression (GTEx) via the Gene Expression Profiling Interactive Analysis (GEPIA2) platform (Tang *et al*., 2019). The platform provided a comprehensive data set of ADC and SCC subtypes, comprising 483 tumors and 347 normal samples in the ADC cohort as well as 486 tumors and 338 normal samples in the SCC cohort. A similar trend was observed, showing lower expression levels of the aforementioned four genes in normal samples as compared to the tumorous samples (**Figure 7**).

**Figure 6.**
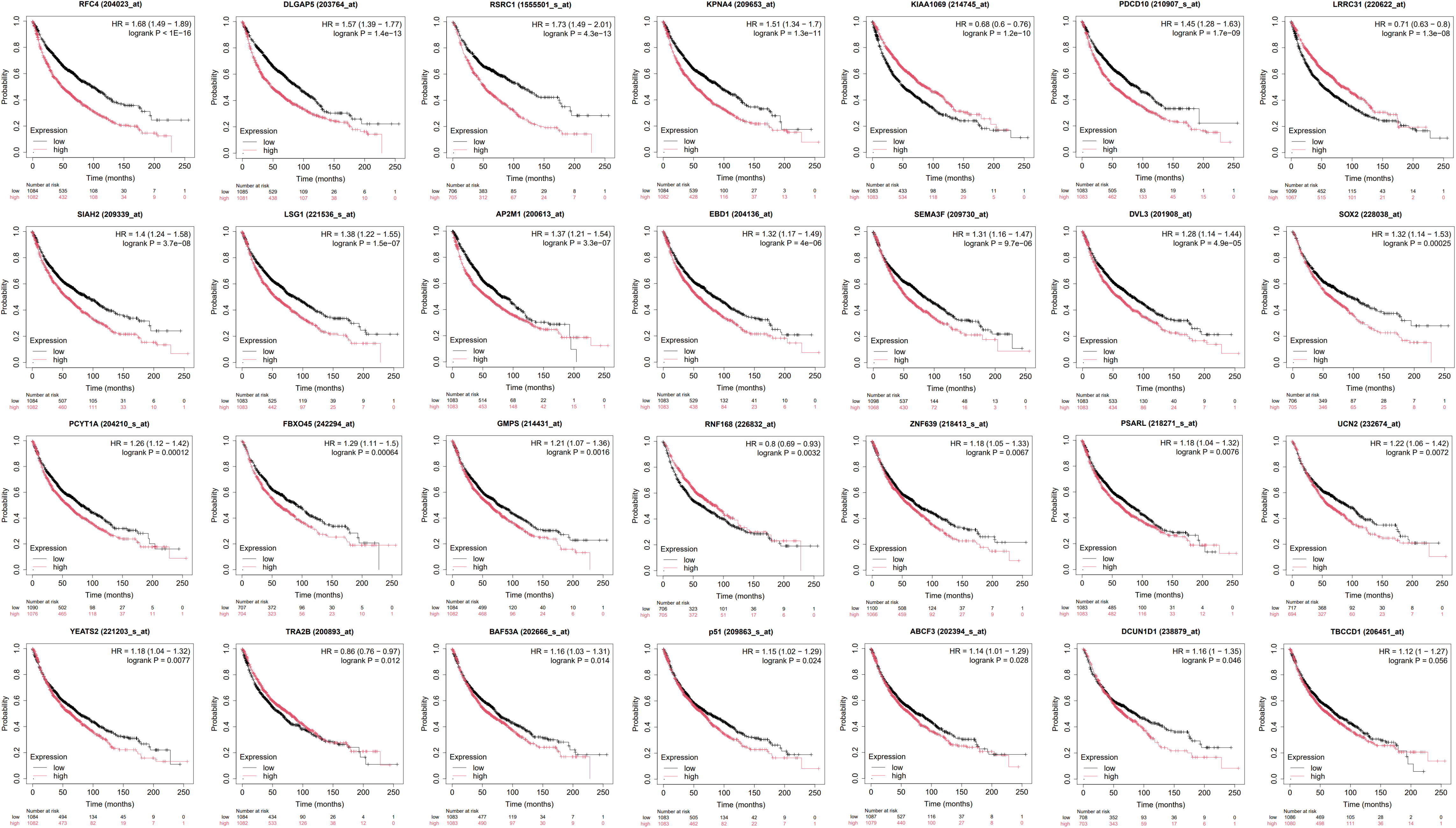
Kaplan-Meier curves of the 28 significant genes (sorted according to p-value) *DLG1*, *PLCH1*, *COL7A1*, *PARL*, *ACTL6A* and *TP63* are aliased as *DLGAP5*, *KIAA1069*, *EBD1*, *PSARL*, *BAF53A* and *P51*, respectively. The x-axis denotes the survival period (months) while the y-axis denotes the probability of survival. The black and red points represent the low and high expression-value of the corresponding gene, respectively. Interestingly, lower expression values of *PLCH1* (*KIAA1069*), *LRRC31*, *RNF168* and *TRA2B* genes are associated with better survival outcomes. In contrast, the remaining 24 genes exhibit an opposite behavior, where the lower expression values correlate to poor prognosis.

**Figure 7.**
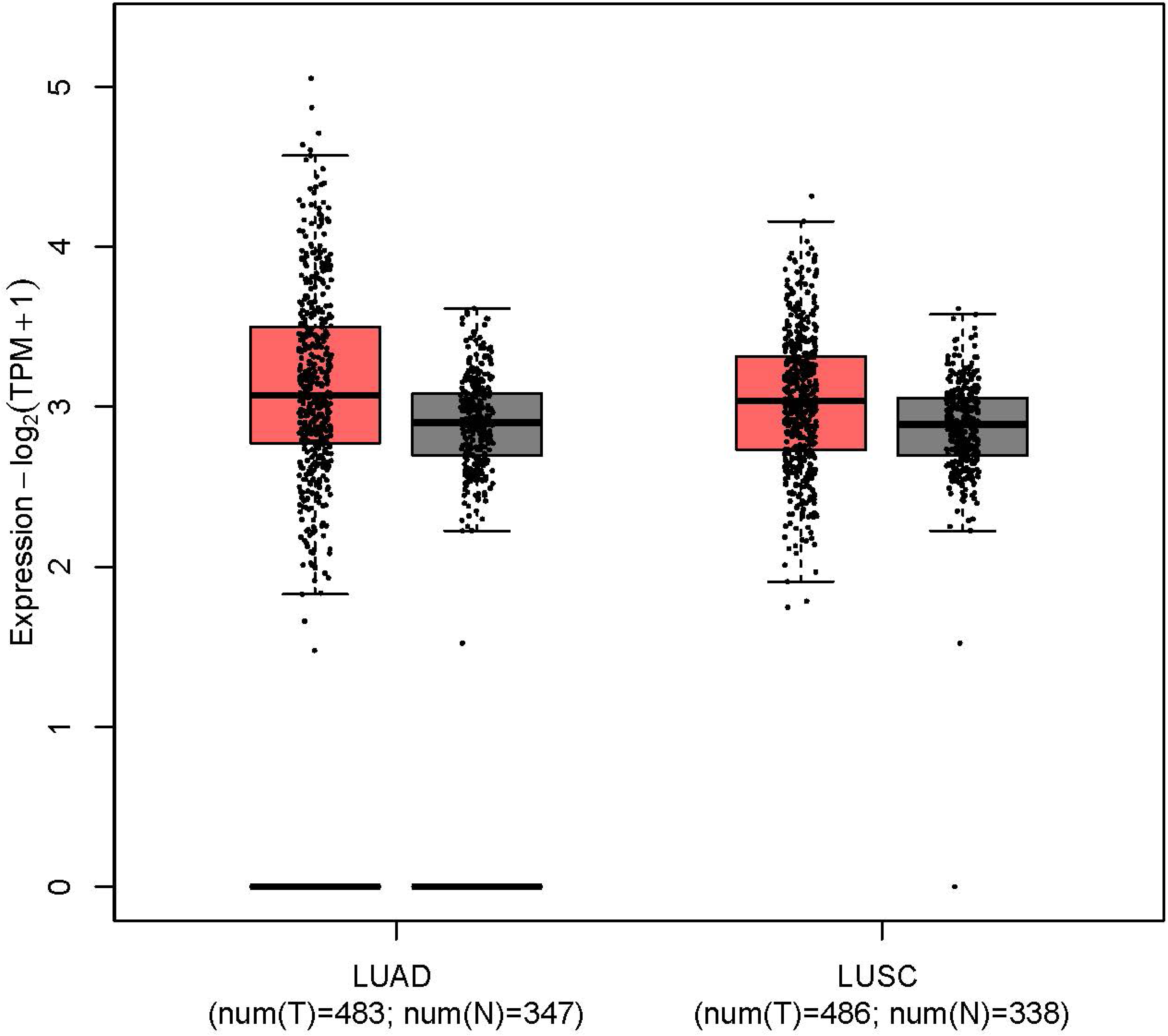
Gene expression-based boxplot of combined expression values of *PLCH1* (*KIAA1069*), *LRRC31*, *RNF168* and *TRA2B* genes. The analysis revealed that the expression of these genes was relatively lower in normal samples as compared to the tumorous samples, thus, supporting their prognostic relevance in the non-small cell lung cancer subtypes lung adenocarcinoma (LUAD) and lung squamous cell carcinoma (LUSC).

## 4 Discussion

The primary objective of the proposed study is to identify a highly accurate approach for the discovery of key NSCLC genes that could assist in improving targeted therapeutic strategies in future. To achieve this, multiple omics data were utilized in an integrative manner to capture the complex interconnected biological dependencies underlying NSCLC subtypes. This multiomics approach to genes identification was implemented by employing a weighted-average-based decision-level fusion mechanism that fused the predicted probabilities generated by ML models trained on individual omics data sets. Initially, the classification performance of individual omics data sets was compared with that of the fusion-based approach by employing various evaluation metrics. As hypothesized, the weighted-average-based decision-level-fusion approach demonstrated superior prediction performance compared to the models trained on individual omics data sets. Thereafter, the identification of a set of NSCLC-relevant genes that are cohesively responsible for accurate subtype classification was performed and unveiled a set of 47 key genes, which play a role in different aspects and stages of NSCLC development (**Figure 4**).

While the publicly available data sets that the study utilized (TCGA repository) are comprehensive in nature, they may introduce inherent biases due to the patient demographics, history of treatment and medication and methodology followed to collect the data (Dwivedi *et al*., 2024a). Further, a key challenge when handling omics data with machine/deep learning techniques is the *curse of dimensionality*. As evident from the employed data (**Table 1**), the cohorts follow high-dimension-low-sample-size (HDLSS) format with instances (patient samples) relatively lower than the number of features (genes). This imbalance may introduce overfitting and lead to reduced generalizability of learned models. In addition, the decision-level fusion technique depends on the predictive strength of the employed individual omics data sets, highlighting the need of incorporating accurate data sets.

In conclusion, the outcome of this work emphasizes that the prediction performance can be enhanced by using a fusion-based multiomics approach, yielding a concise set of disease-relevant genes. Future work could enhance the framework by incorporating more omics-level data sets together with histopathological images of the NSCLC samples. Of course, this approach could also be applied to any other type of disease and could be of particular interest for those pathologies that currently lack a funded understanding of their underlying molecular mechanisms. This multi-modal approach may improve overall classification performance and assist in a more robust identification of molecular targets which will have to be evaluated biologically towards determining their potential as targets in diagnosis and treatment strategies.

## Data Availability

All data produced in the present study are available upon reasonable request to the authors.

## Acknowledgements

The authors would like to thank Akshat Kishore Srivastava, Department of Electrical Engineering and Computer Science, University of California, Berkeley, Berkeley, California, 94720, USA, for his valuable discussions on machine learning model development.

## References

Arik, S.Ö. and Pfister, T. (2021) ‘Tabnet: Attentive interpretable tabular learning’, in Proceedings of the AAAI conference on artificial intelligence, pp. 6679–6687.

Bolón-Canedo, V., Sánchez-Maroño, N. and Alonso-Betanzos, A. (2013) ‘A review of feature selection methods on synthetic data’, Knowledge and information systems, 34, pp. 483–519.

Breiman, L. (2001) ‘Random forests’, Machine learning, 45, pp. 5–32.

Cai, Z. et al. (2015) ‘Classification of lung cancer using ensemble-based feature selection and machine learning methods’, Molecular BioSystems, 11(3), pp. 791–800.

Carrillo-Perez, F. et al. (2022) ‘Machine-learning-based late fusion on multi-omics and multi-scale data for non-small-cell lung cancer diagnosis’, Journal of Personalized Medicine, 12(4), p. 601.

Chen, J.W. and Dhahbi, J. (2021) ‘Lung adenocarcinoma and lung squamous cell carcinoma cancer classification, biomarker identification, and gene expression analysis using overlapping feature selection methods’, Scientific reports, 11(1), p. 13323.

Chen, R.J. et al. (2022) ‘Pan-cancer integrative histology-genomic analysis via multimodal deep learning’, Cancer Cell, 40(8), pp. 865–878.

Chen, T. and Guestrin, C. (2016) ‘Xgboost: A scalable tree boosting system’, in Proceedings of the 22nd acm sigkdd international conference on knowledge discovery and data mining, pp. 785–794.

Cortes, C. and Vapnik, V. (1995) ‘Support-vector networks’, Machine learning, 20, pp. 273–297.

Denu, R.A. and Burkard, M.E. (2020) ‘Analysis of the “centrosome-ome” identifies MCPH1 deletion as a cause of centrosome amplification in human cancer’, Scientific reports, 10(1), p. 11921.

Dobin, A. et al. (2013) ‘STAR: ultrafast universal RNA-seq aligner’, Bioinformatics, 29(1), pp. 15– 21.

Dwivedi, K. et al. (2023) ‘An explainable AI-driven biomarker discovery framework for Non-Small Cell Lung Cancer classification’, Computers in Biology and Medicine, 153, p. 106544.

Dwivedi, K. et al. (2024a) ‘Enlightening the path to NSCLC biomarkers: Utilizing the power of XAI-guided deep learning’, Computer Methods and Programs in Biomedicine, 243, p. 107864.

Dwivedi, K. et al. (2024b) ‘XL1R-Net: Explainable AI-driven improved L1-regularized deep neural architecture for NSCLC biomarker identification’, Computational Biology and Chemistry, 108, p. 107990.

Fabregat, A. et al. (2017) ‘Reactome pathway analysis: a high-performance in-memory approach’, BMC bioinformatics, 18, pp. 1–9.

Fan, J. et al. (2023) ‘ABC transporters affects tumor immune microenvironment to regulate cancer immunotherapy and multidrug resistance’, Drug Resistance Updates, 66, p. 100905.

Fix, E. and Hodges, J.L. (1989) ‘Discriminatory analysis. Nonparametric discrimination: Consistency properties’, International Statistical Review/Revue Internationale de Statistique, 57(3), pp. 238–247.

Girard, L. et al. (2016) ‘An expression signature as an aid to the histologic classification of non– small cell lung cancer’, Clinical Cancer Research, 22(19), pp. 4880–4889.

Goldman, M.J. et al. (2020a) ‘Visualizing and interpreting cancer genomics data via the Xena platform’, Nature biotechnology, 38(6), pp. 675–678.

Goldman, M.J. et al. (2020b) ‘Visualizing and interpreting cancer genomics data via the Xena platform’, Nature biotechnology, 38(6), pp. 675–678.

Guinney, J. et al. (2015) ‘The consensus molecular subtypes of colorectal cancer’, Nature medicine, 21(11), pp. 1350–1356.

Györffy, B. et al. (2010) ‘An online survival analysis tool to rapidly assess the effect of 22,277 genes on breast cancer prognosis using microarray data of 1,809 patients’, Breast cancer research and treatment, 123, pp. 725–731.

Hao, Z. et al. (2008) ‘Urocortin2 inhibits tumor growth via effects on vascularization and cell proliferation’, Proceedings of the National Academy of Sciences, 105(10), pp. 3939–3944.

Haykin, S. (1994) Neural networks: a comprehensive foundation. Prentice Hall PTR.

Hoffman, P. et al. (1997) ‘DNA visual and analytic data mining’, in Proceedings. Visualization’97 (Cat. No. 97CB36155). IEEE, pp. 437–441.

Imoto, I. et al. (2003) ‘Identification of ZASC1 encoding a Kruppel-like zinc finger protein as a novel target for 3q26 amplification in esophageal squamous cell carcinomas’, Cancer research, 63(18), pp. 5691–5696.

Kanehisa, M. and Goto, S. (2000) ‘KEGG: kyoto encyclopedia of genes and genomes’, Nucleic acids research, 28(1), pp. 27–30.

Karlos, S., Kostopoulos, G. and Kotsiantis, S. (2020) ‘A soft-voting ensemble based co-training scheme using static selection for binary classification problems’, Algorithms, 13(1), p. 26.

Khadirnaikar, S., Shukla, S. and Prasanna, S. (2023) ‘Machine learning based combination of multi-omics data for subgroup identification in non-small cell lung cancer’, Scientific Reports, 13(1), p. 4636.

Kim, T.-M. et al. (2013) ‘Functional genomic analysis of chromosomal aberrations in a compendium of 8000 cancer genomes’, Genome research, 23(2), pp. 217–227.

Kononenko, I. (1994) ‘Estimating attributes: Analysis and extensions of RELIEF’, in European conference on machine learning. Springer, pp. 171–182.

Kononenko, I., Šimec, E. and Robnik-Šikonja, M. (1997) ‘Overcoming the myopia of inductive learning algorithms with RELIEFF’, Applied Intelligence, 7, pp. 39–55.

Lánczky, A. and Gy\Horffy, B. (2021) ‘Web-based survival analysis tool tailored for medical research (KMplot): development and implementation’, Journal of medical Internet research, 23(7), p. e27633.

Li, B.-Q. et al. (2014) ‘Classification of non-small cell lung cancer based on copy number alterations’, PLoS One, 9(2), p. e88300.

Lipkova, J. et al. (2022) ‘Artificial intelligence for multimodal data integration in oncology’, Cancer cell, 40(10), pp. 1095–1110.

Liu, H. and Setiono, R. (1998) ‘Incremental feature selection’, Applied Intelligence, 9, pp. 217– 230.

Liu, Y. et al. (2022) ‘Circular RNA circACAP2 Suppresses Ferroptosis of Cervical Cancer during Malignant Progression by miR-193a-5p/GPX4’, Journal of oncology, 2022(1), p. 5228874.

Lundberg, S.M. and Lee, S.-I. (2017) ‘A unified approach to interpreting model predictions’, CoRR, abs/1705.07874. Available at: http://arxiv.org/abs/1705.07874.

McClish, D.K. (1989) ‘Analyzing a portion of the ROC curve’, Medical decision making, 9(3), pp. 190–195.

Mermel, C.H. et al. (2011) ‘GISTIC2. 0 facilitates sensitive and confident localization of the targets of focal somatic copy-number alteration in human cancers’, Genome biology, 12, pp. 1–14.

Mi, H. and Thomas, P. (2009) ‘PANTHER pathway: an ontology-based pathway database coupled with data analysis tools’, in Protein networks and pathway analysis. Springer, pp. 123–140.

Morrison, J.L. et al. (2005) ‘GeneRank: using search engine technology for the analysis of microarray experiments’, BMC bioinformatics, 6, pp. 1–14.

Olshen, A.B. et al. (2004) ‘Circular binary segmentation for the analysis of array-based DNA copy number data’, Biostatistics, 5(4), pp. 557–572.

Peng, H., Long, F. and Ding, C. (2005) ‘Feature selection based on mutual information criteria of max-dependency, max-relevance, and min-redundancy’, IEEE Transactions on pattern analysis and machine intelligence, 27(8), pp. 1226–1238.

Pfarr, N. et al. (2016) ‘Copy number changes of clinically actionable genes in melanoma, non-small cell lung cancer and colorectal cancer—A survey across 822 routine diagnostic cases’, Genes, Chromosomes and Cancer, 55(11), pp. 821–833.

Pinkel, D. and Albertson, D.G. (2005) ‘Array comparative genomic hybridization and its applications in cancer’, Nature genetics, 37(Suppl 6), pp. S11–S17.

Qi, L. et al. (2021) ‘Multi-omics data fusion for cancer molecular subtyping using sparse canonical correlation analysis’, Frontiers in genetics, 12, p. 607817.

Qiu, Z.-W. et al. (2017) ‘Genome-wide copy number variation pattern analysis and a classification signature for non-small cell lung cancer’, Genes, Chromosomes and Cancer, 56(7), pp. 559–569.

Rastghalam, R. and Pourghassem, H. (2016) ‘Breast cancer detection using MRF-based probable texture feature and decision-level fusion-based classification using HMM on thermography images’, Pattern Recognition, 51, pp. 176–186.

Shapley, L.S. and others (1953) ‘A value for n-person games’.

Shen, D. et al. (2022) ‘METTL14-mediated Lnc-LSG1 m6A modification inhibits clear cell renal cell carcinoma metastasis via regulating ESRP2 ubiquitination’, Molecular Therapy Nucleic Acids, 27, pp. 547–561.

Siegel, R.L., et al. (2022) ‘Cancer statistics, 2022’, CA: A Cancer Journal for Clinicians, 72(1), pp. 7–33. Available at: 10.3322/caac.21708.

Steyaert, S. et al. (2023) ‘Multimodal data fusion for cancer biomarker discovery with deep learning’, Nature machine intelligence, 5(4), pp. 351–362.

Tang, Z. et al. (2019) ‘GEPIA2: an enhanced web server for large-scale expression profiling and interactive analysis’, Nucleic acids research, 47(W1), pp. W556–W560.

Thomas, P.D. et al. (2022) ‘PANTHER: Making genome-scale phylogenetics accessible to all’, Protein Science, 31(1), pp. 8–22.

Tian, S. (2017) ‘Classification and survival prediction for early-stage lung adenocarcinoma and squamous cell carcinoma patients’, Oncology letters, 14(5), pp. 5464–5470.

Travis, W.D. et al. (2015) ‘The 2015 World Health Organization classification of lung tumors: impact of genetic, clinical and radiologic advances since the 2004 classification’, Journal of thoracic oncology, 10(9), pp. 1243–1260.

Urbanowicz, R.J. et al. (2018) ‘Benchmarking relief-based feature selection methods for bioinformatics data mining’, Journal of biomedical informatics, 85, pp. 168–188.

Weissmiller, A.M., Fesik, S.W. and Tansey, W.P. (2024) ‘WD repeat domain 5 inhibitors for cancer therapy: not what you think’, Journal of Clinical Medicine, 13(1), p. 274.

Yang, Y. et al. (2022) ‘MDICC: novel method for multi-omics data integration and cancer subtype identification’, Briefings in Bioinformatics, 23(3), p. bbac132.

Zhang, B., Kirov, S. and Snoddy, J. (2005) ‘WebGestalt: an integrated system for exploring gene sets in various biological contexts’, Nucleic acids research, 33(suppl_2), pp. W741–W748.

